# The Impact of the “Muslim Ban” Executive Order on Healthcare Utilization in Minneapolis-St. Paul, Minnesota

**DOI:** 10.1101/2020.10.23.20218628

**Authors:** Elizabeth A. Samuels, Lilla Orr, Elizabeth B. White, Altaf Saadi, Aasim I. Padela, Michael Westerhaus, Aarti D. Bhatt, Pooja Agrawal, Dennis Wang, Gregg Gonsalves

## Abstract

**Objective:** Determine whether the 2017 “Muslim Ban” Executive Order impacted healthcare utilization by people born in Order-targeted nations living in the United States.

**Methods:** We conducted a retrospective cohort study of people living in Minneapolis-St. Paul, MN in 2016-2017 who were: 1) born in Order-targeted nations, 2) born in Muslim-majority nations not listed in the Order, and 3) born in the United States and non-Latinx. Primary outcomes were: 1) primary care visits, 2) missed primary care appointments, 3) primary care diagnoses for stress-responsive conditions, 4) emergency department visits, and 5) emergency department visits for stress-responsive diagnoses. We evaluated visit trends before and after Order issuance using linear regression and differences between study groups using a difference-in-difference analyses.

**Results:** In early 2016, primary care visits and stress-responsive diagnoses increased among individuals from Muslim majority nations. Following the Order, there was an immediate increase in emergency department visits among individuals from Order-targeted nations.

**Conclusions:** Increases in healthcare utilization among people born in Muslim majority countries before and after the “Muslim Ban” likely reflect elevated cumulative stress including the impact of the Order.

## INTRODUCTION

The 2016 United States (U.S.) presidential election was marked by anti-Muslim and anti-immigrant rhetoric and the subsequent Trump administration has introduced multiple restrictive immigration policies, primarily targeting individuals from Muslim-majority and Latin American countries.^1^ On January 27, 2017, one week after taking office, President Trump issued Executive Order 13769, “Protecting the Nation from Foreign Terrorist Entry into the United States,”^2^ commonly referred to as the “Muslim Ban.” The Order, upheld by the U.S. Supreme Court,^3^ suspended the U.S. Refugee Resettlement Program and prevented citizens from seven Muslim-majority countries (Iraq, Syria, Iran, Libya, Somalia, Sudan, and Yemen) from traveling or immigrating to the U.S.

Policies like the Muslim Ban exacerbate heightened levels of discrimination, hostility, and “othering” that U.S. Muslims experience.^4^ Over the past two decades there has been an increase in hate crimes^5^ and social hostility^6,7^ directed toward U.S. Muslims, experiences which negatively impact health. Following the September 11^th^ attacks, Arab Americans, including Muslim Arab Americans, demonstrated increased rates of anxiety, depression, and low birth weights.^4,8-11^ However, it is unknown how health and healthcare utilization in other Muslim American communities have changed in response to shifting sociopolitical climates.

This study examines the impact of the Muslim Ban Order on healthcare utilization among people from Order-targeted nations. We sought to determine whether the policy resulted in avoidance of care due to fear of discrimination, or, as was seen among Arab Americans after September 11^th^, increased healthcare utilization for stress-responsive medical problems. To evaluate these changes, we examined primary care and emergency department (ED) utilization by people from Order-targeted nations living in the Minneapolis-St. Paul, Minnesota metropolitan area, home to the largest Somali Muslim community in the U.S.^12^

## METHODS

### Study Design

We conducted a retrospective cohort study comparing changes in primary care and ED utilization, missed scheduled clinic appointments, and visits for stress-responsive conditions among individuals from Order-targeted nations from one year before to one year after Order issuance. We characterized visit trends and, for outcomes with similar group trends prior to Order issuance, used a difference-in-difference analysis to estimate its effects on healthcare utilization. The primary analysis compared people born in one of the Order-targeted nations to non-Latinx U.S.-born citizens. Supplementary analyses compared trends among people born in a Muslim-majority nation not listed in the Order (Table 1, Group 2) to non-Latinx U.S.-born citizens.

**Table 1.**
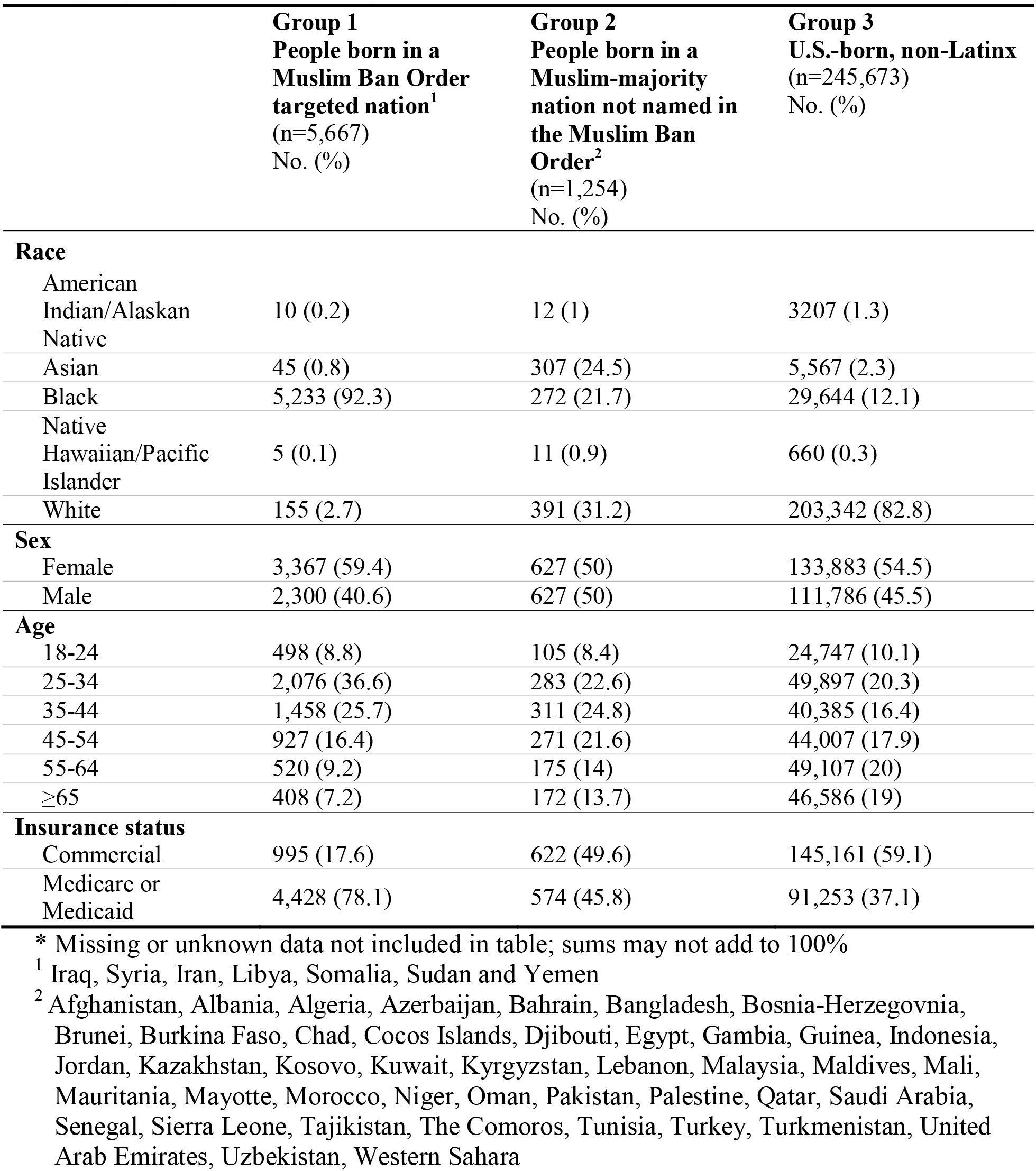
Characteristics of HealthPartners patients seeking care in a primary care clinic or emergency department, January 2016 to December 2017.

|  | **Group 1**<br>**People born in a**<br>**Muslim Ban Order**<br>**targeted nation<sup>1</sup>**<br>**(n=5,667)**<br>**No. (%)** | **Group 2**<br>**People born in a**<br>**Muslim-majority**<br>**nation not named in**<br>**the Muslim Ban**<br>**Order<sup>2</sup>**<br>**(n=1,254)**<br>**No. (%)** | **Group 3**<br>**U.S.-born, non-Latinx**<br>**(n=245,673)**<br>**No. (%)** |
| --- | --- | --- | --- |
| **Race** |  |  |  |
| American<br>Indian/Alaskan<br>Native | 10 (0.2) | 12 (1) | 3207 (1.3) |
| Asian | 45 (0.8) | 307 (24.5) | 5,567 (2.3) |
| Black | 5,233 (92.3) | 272 (21.7) | 29,644 (12.1) |
| Native<br>Hawaiian/Pacific<br>Islander | 5 (0.1) | 11 (0.9) | 660 (0.3) |
| White | 155 (2.7) | 391 (31.2) | 203,342 (82.8) |
| **Sex** |  |  |  |
| Female | 3,367 (59.4) | 627 (50) | 133,883 (54.5) |
| Male | 2,300 (40.6) | 627 (50) | 111,786 (45.5) |
| **Age** |  |  |  |
| 18-24 | 498 (8.8) | 105 (8.4) | 24,747 (10.1) |
| 25-34 | 2,076 (36.6) | 283 (22.6) | 49,897 (20.3) |
| 35-44 | 1,458 (25.7) | 311 (24.8) | 40,385 (16.4) |
| 45-54 | 927 (16.4) | 271 (21.6) | 44,007 (17.9) |
| 55-64 | 520 (9.2) | 175 (14) | 49,107 (20) |
| ≥65 | 408 (7.2) | 172 (13.7) | 46,586 (19) |
| **Insurance status** |  |  |  |
| Commercial | 995 (17.6) | 622 (49.6) | 145,161 (59.1) |
| Medicare or<br>Medicaid | 4,428 (78.1) | 574 (45.8) | 91,253 (37.1) |

\* Missing or unknown data not included in table; sums may not add to 100%
<sup>1</sup> Iraq, Syria, Iran, Libya, Somalia, Sudan and Yemen
<sup>2</sup> Afghanistan, Albania, Algeria, Azerbaijan, Bahrain, Bangladesh, Bosnia-Herzegovnia, Brunei, Burkina Faso, Chad, Cocos Islands, Djibouti, Egypt, Gambia, Guinea, Indonesia, Jordan, Kazakhstan, Kosovo, Kuwait, Kyrgyzstan, Lebanon, Malaysia, Maldives, Mali, Mauritania, Mayotte, Morocco, Niger, Oman, Pakistan, Palestine, Qatar, Saudi Arabia, Senegal, Sierra Leone, Tajikistan, The Comoros, Tunisia, Turkey, Turkmenistan, United Arab Emirates, Uzbekistan, Western Sahara

### Study setting and population

The Minneapolis-St. Paul metropolitan area has 3.63 million residents and the largest Somali Muslim population in the U.S. In 2017, three quarters (75.1%) of Minneapolis-St. Paul metropolitan area residents were white, 8.56% Black, and 6.64% Asian.^13^ Approximately 10.2% of residents were born outside of the U.S., with an estimated 37,468 people born in Somalia.^13^

We analyzed electronic health record (EHR) data from HealthPartners, one of the area’s largest healthcare and insurance organizations, serving over 1.2 million patients at 55 primary care centers, 22 acute care centers, and eight hospitals in the Minneapolis-St. Paul metropolitan area. While religion is not recorded in the EHR, patient country of origin recorded in the HealthPartners EHR allowed us to characterize patients receiving care between January 1, 2016 and December 31, 2017 into three groups: 1) adults born in one of the seven nations mentioned in the Muslim Ban Order (Iran, Iraq, Libya, Somalia, Sudan, Syria, and Yemen) (Table 1, Group 1), 2) adults born in Muslim-majority nations not listed in the Order (Table 1, Group 2), and 3) U.S.-born non-Latinx adults (Table 1, Group 3). We excluded U.S.-born Latinx patients as they have been subject to distinct anti-immigrant rhetoric and policies which have important impacts on their health and healthcare utilization.^14,15^

### Outcomes

We examined changes in primary care and ED utilization in the year before and after Order issuance. Primary outcomes included the number of 1) primary care clinic visits, 2) missed primary care clinic appointments, 3) primary care clinic diagnoses for stress-responsive conditions, 4) ED visits, and 5) ED visits for stress-responsive diagnoses, including ambulatory sensitive conditions. This study was conducted in accordance with STROBE guidelines and approved by the HealthPartners and Yale Institutional Review Boards.^16^

#### Primary Care Clinic Utilization

Primary care visits, missed appointments, and stress-responsive diagnoses were analyzed as counts per person. We identified stress-responsive diagnoses, medical diagnoses that may be related to increased stress, through literature review and expert opinion.^10,14,17-23^ Diagnoses included in the analysis were agreed upon by consensus (Table S1) and included 138 ICD-10 codes grouped into six categories: mental health, sleep disorders, gastrointestinal concerns, neurologic concerns, food-related disorders, and pain syndromes.

#### Emergency Department Utilization

ED visits, stress-responsive diagnoses, and ambulatory sensitive conditions were also analyzed as counts per person. We identified ED stress-responsive diagnoses through literature review and expert opinion. Diagnoses included were agreed upon by research team consensus and included 27 ICD-10 codes for acute coronary syndrome, assault, suicide attempt, and syncope (Table S2).^10,14,17-25^ Ambulatory sensitive conditions are conditions responsive to social stressors and inequalities for which an ED visit or hospitalization is considered preventable through outpatient interventions.^24,26^ Ambulatory sensitive diagnoses included 21 ICD-10 codes for: angina, asthma, congestive heart failure, chronic obstructive pulmonary disease, diabetes complications, and hypertension (Table S2).^24,26^

### Data

All patient demographic, visit, and diagnosis data were extracted from the HealthPartners EHR by a HealthPartners Data Analyst. All records were de-identified and assigned a unique Study ID prior to transfer through a secure file transmission system from HealthPartners to the study team.

### Statistical Analyses

We summarized the trends for each outcome using local linear regression and tested trend similarity across groups prior to Order issuance using linear regression. For the difference-in-difference analyses, we fit the linear regression model described in equation 1.

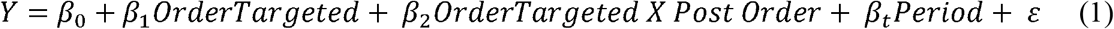

We compared the 360-days before and after Order issuance, divided into 24 distinct, 30-day time periods. Each outcome “Y” is a count per person per 30-day time period. “Order Targeted” and “Order Targeted x Post Order” represent being from a nation named in the Order and being from one of these nations in a time period after the Order was issued, respectively. For each model, we also estimated the average effect (*β*_2_) over increasing time intervals centered on the Order issuance date, beginning with 30-days pre- and post-Order and increasing by 30-day increments to 360-days pre- and post-Order.

To further control for potentially confounding effects of individual characteristics and temporal trends, we estimated the average effects relative to two alternative reference groups. First, we used exact matching on age, sex, race, and insurance to reweight members of Group 3 (non-Latinx, U.S. born) and identify a subset of Group 3 with similar demographics as Group 1 (Order-targeted). We used the R package MatchIt to identify this reference group, then fit a weighted version of the model described in equation 1 (Table S8).^27^ Second, we used a generalized synthetical control method to reweight members of Group 3 to produce a reference group with demographics and pre-Order outcomes more similar to those observed in Group 1. Models were fit using the R package gsynth with parametric bootstrap standard errors.^28^

We fit separate models for all outcomes: primary care visits, missed primary care appointments, primary care stress-responsive diagnoses, ED visits, and ED stress-responsive diagnoses. The primary difference-in-difference analyses estimated the change in outcomes between pre- and post-Order periods among individuals from Order-targeted nations above and beyond the change observed among non-Latinx U.S.-born individuals. To determine whether these differences are due to the Order, rather than other time-varying differences, we assumed that the trends would have been equivalent across groups if the order had not been issued. We tested for parallel trends in the pre-intervention period by examining the interaction between study group and time (30-day periods) in linear regression models. Our main results include difference-in-difference estimates of the effect of the Order for outcomes with outcomes that did not violate the parallel trends assumption prior to Order issuance. All estimates and robustness checks are reported in the Supplement.

## RESULTS

### Characteristics of Study Population

From 2016 to 2017, 252,594 patients were included in this analysis: 5,667 (2.2%) in Group 1, 1,254 (0·5%) in Group 2, and 245,673 (97.3%) in Group 3 (Table 1). People in Group 1 (from Order-targeted nations) were predominantly Black (92.3%, 5,233/5,667), female (59·4%, 3,367/5,667), 25-44 years of age (62.3%, 3,534/5,667), and had Medicare or Medicaid (78.1%, 4,428/5,667). The largest proportion of Group 2 (non-Order-targeted Muslim-majority nations) identified as white (31.2%, 391/1,254) and 22-54 years of age (69%, 865/1,254). Group 3 (U.S.- born, non-Latinx) was predominantly white (82.8%, 203,342/245,673), slightly more than half were female (54.5% 133,883/245,673), and had an older age distribution (Table 1).

### Trends in healthcare utilization and diagnoses

The Muslim Ban Order was issued during a period of political change and its implementation did not occur on a single day. Characterizing visit and diagnosis trends is critical to understanding the effects of the political environment during 2016 and 2017 and allows us to assess for parallel trends prior to Order issuance. Figure 1 displays weekly average visit counts per person for each group, along with a local linear (loess) approximation of the time trend.

**Figure 1:**
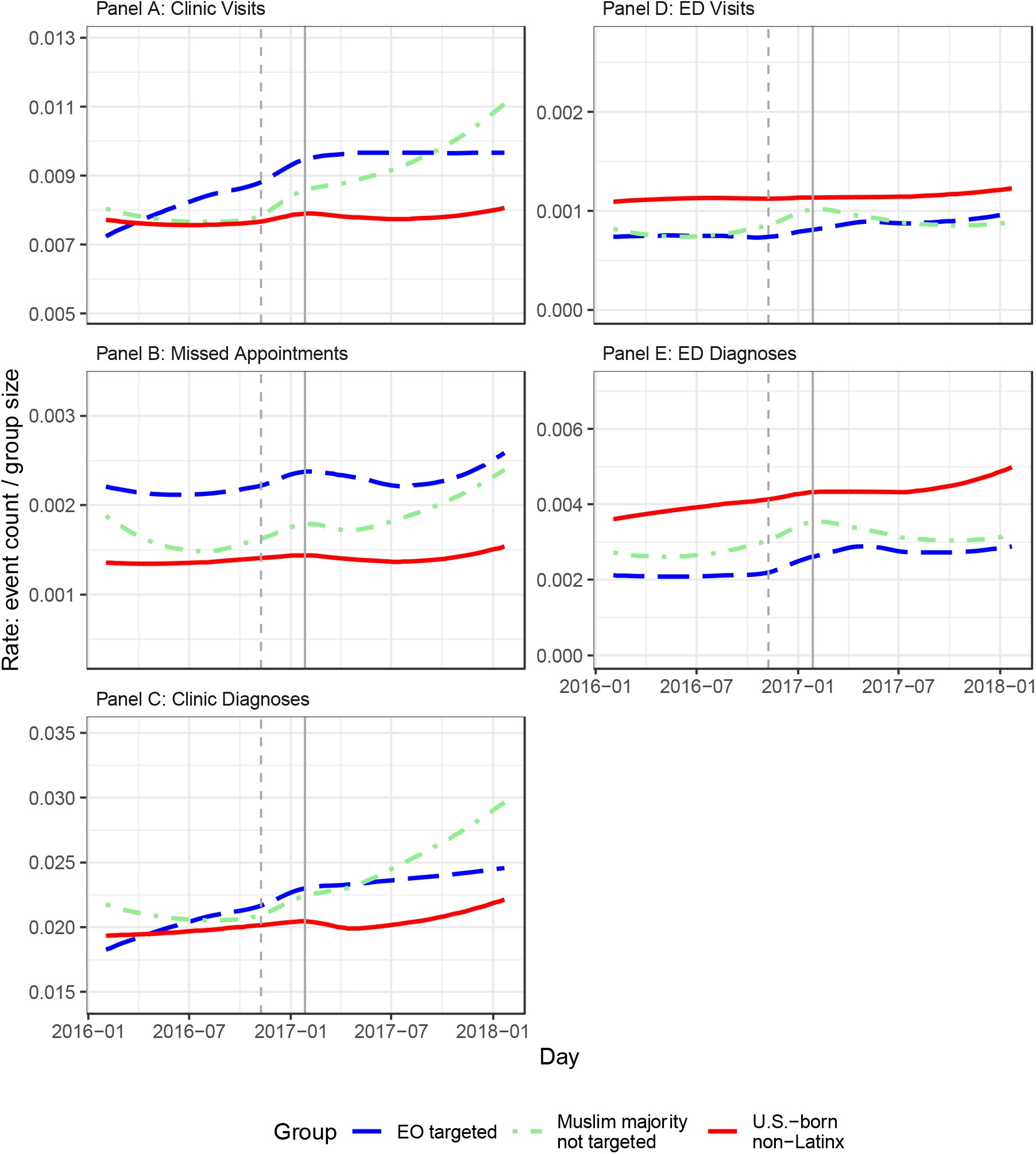
Time trends for all primary outcomes among patients from Order-targeted nations, patients from other Muslim-majority nations, and U.S.-born non-Latinx patients, January 2016 to December 2017. Points indicate weekly average counts per person in each group for A) clinic visits, B) missed clinic appointments, C) clinic stress-responsive diagnoses, D) ED visits, and E) ED stress-responsive diagnoses. A loess regression line summarizing the time trend is included for each group, based on daily average counts per person. For all clinic outcomes, panels A, B, and C, non-business days are excluded from the analysis. The solid line marks the Order issuance and the dotted line marks the 2016 election, for reference.

#### Primary Care Clinic Utilization

Average daily clinic visits and stress-responsive diagnoses trends are similar across all three groups in early 2016 (Figure 1A, 1C). While visits and stress-responsive diagnoses remained fairly constant for U.S.-born non-Latinx individuals (Group 3), beginning in early 2016, they dramatically increased for individuals from Muslim majority nations in both Groups 1 and 2 after the 2016 presidential election and before Order issuance (Figure 1A, 1C). This increase, and therefore the absence of parallel trends in the pre-Order period, means that difference-in-difference analysis cannot be used to identify the effects of the Order on primary care visits and responsive diagnoses (Table S3, S4). Missed scheduled clinic appointments do appear to follow parallel trends prior to Order issuance (Table S3, S4), and difference-in-difference can be conducted.

#### Emergency Department Utilization

U.S.-born non-Latinx individuals have higher baseline ED utilization and ED visits for stress-responsive diagnoses; however, trends are fairly similar for all three groups prior to Order issuance. The rate of ED visits was mostly flat, while stress-responsive diagnoses slightly increased during the year prior to the Order. Around the 2016 election, the rate of ED visits and stress-responsive diagnoses increased for individuals from Order-targeted nations (Group 1) as well as individuals from other Muslim majority nations (Group 2) before leveling off at a higher utilization rate in mid to late 2017.

#### Effect of the Muslim Ban Order on healthcare utilization and diagnoses

For the three outcomes with similar trends between study groups prior to Order issuance, we used difference-in-difference analysis to estimate the effect of the Order (Table 2, Figure 2). Equivalent analysis is presented in the Supplement for all outcomes without parallel trends prior to Order issuance.

**Table 2:**
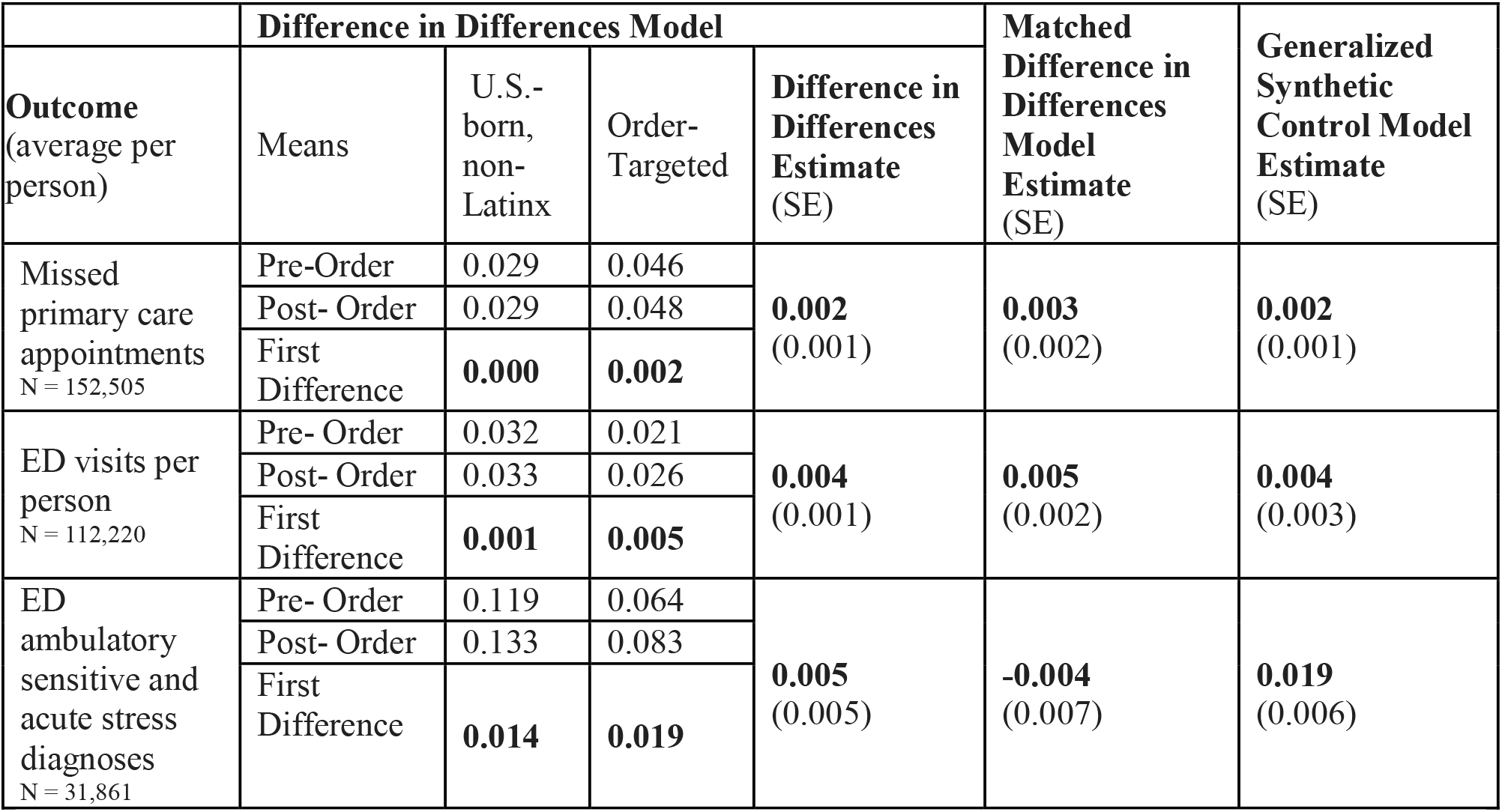
Difference-in-difference estimates of the effect of the Muslim Ban Order on missed primary care clinic appointments, emergency department visits, and emergency department stress-responsive diagnoses among patients from Order-targeted nations (Group 1) Effect estimates are additional increases in each outcome (per person per 30-day time period) from the year before to the year after the Muslim Ban Order was issued among individuals from Order-targeted nations, beyond the increases observed in a reference group. Each outcome is displayed on a separate row. Robust standard errors are included in parentheses for difference in difference estimates with and without demographic matching. Parametric bootstrap standard errors are included in parentheses for generalized synthetical control model estimates. See Supplement for details.

**Figure 2:**
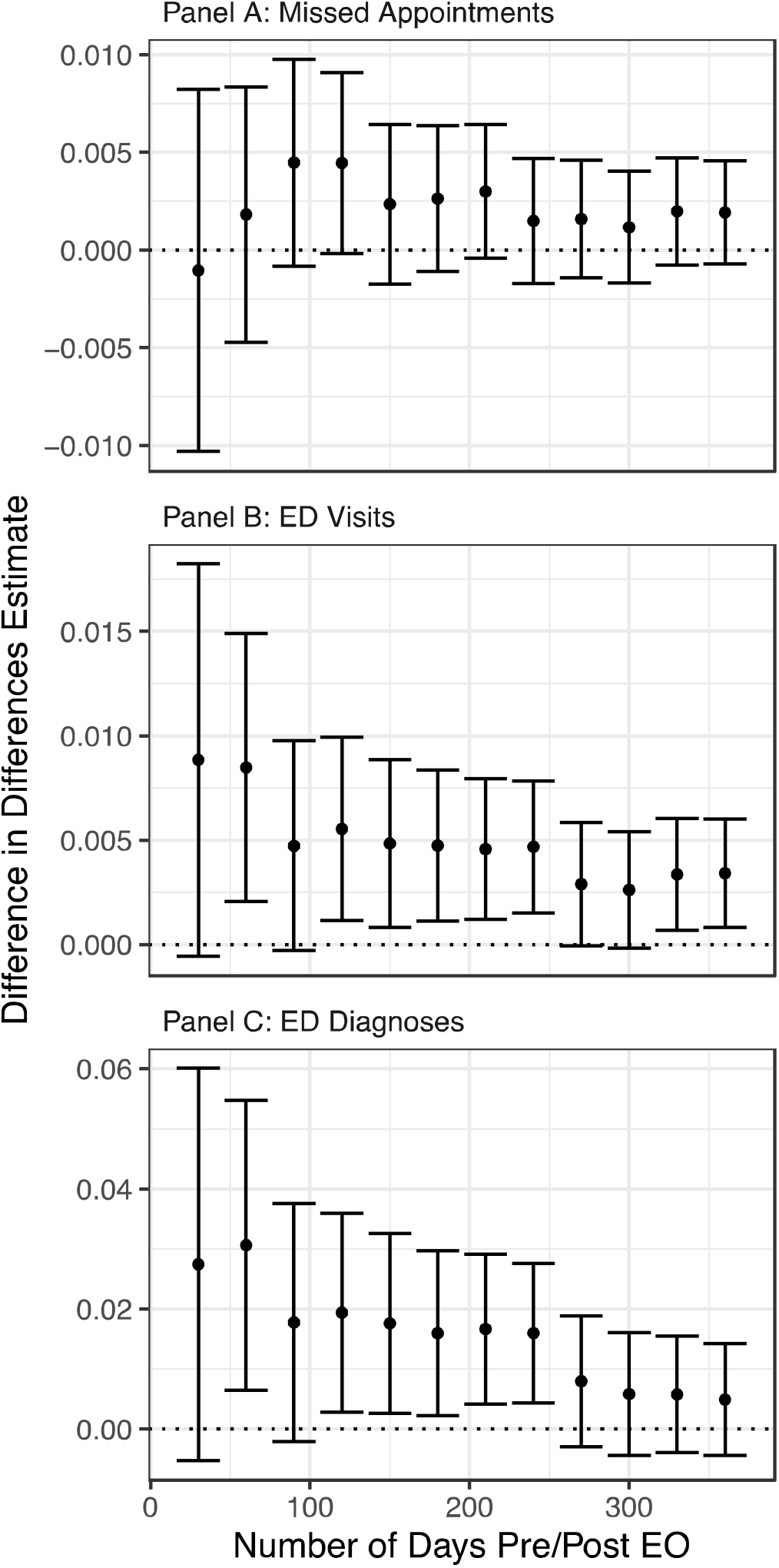
Difference-in-difference estimates for missed clinic appointments, ED visits, and ED stress-responsive diagnoses, January 2016 to December 2017. Each point represents a difference-in-differences estimate of the effect of the Muslim Ban Order on A) missed appointments, B) ED visits, or C) stress-responsive ED diagnoses, with a 95% confidence interval. The left most points compare the difference in each outcome 30 days before to 30 days after the issuance of the Order for Groups 1 and 3. Each additional point compares differences across a larger time period up to 360 days pre-/post-Order.

#### Primary Care Clinic Utilization

In the year following issuance of the Muslim Ban Order, Groups 1 and 2 had greater increases in rates of primary care visits than were seen in Group 3 or either alternative reference group drawn from Group 3 (Tables S5, S7, S9). However, the rise in clinic visits and stress-responsive diagnoses occurred before Order issuance. Rates of missed appointments for Group 1 do not appear to have been affected by the Order (Table 2, Figure 2A), although a slight positive effect is estimated for Group 2 (Table S14).

#### Emergency Department Utilization

In the year following Order issuance, there was an increase in ED visits and ED stress-responsive diagnoses among individuals from Order-targeted nations beyond the increase seen among U.S.-born non-Latinx individuals. (Table 2, Figure 2B). There was an additional increase in ED stress-responsive diagnoses after issuance of the Order for individuals from Order-targeted nations; however, the difference-in-differences estimate was not statistically significant. Compared to the demographically matched reference group, the estimated Order effect on the Order-targeted group was negative, but also not statistically significant (Table 2, S7). Compared to the synthetic control, the estimate was larger and statistically significant, but the confidence intervals overlap. These findings are consistent, but together they do not provide strong evidence of an Order effect on stress-responsive ED diagnoses in the year following the order. (Table 2). The estimated effects of the Muslim Ban Order on ED utilization and stress-responsive diagnoses for individuals from nations not targeted in the Order were also not statistically significant (Table S14).

In contrast to the utilization trends in the primary care clinic, Group 1 ED visits and stress-responsive diagnoses may have been more immediately impacted by the Muslim Ban Order. Shortly after the Order is issued, in the first 30- to 60-days, difference-in-difference analysis demonstrates large Order effects on ED visits (Figure 2B) and stress-responsive diagnoses (Figure 2C). Both increase shortly after the Order was issued beyond the increase seen in Group 3, but is not statistically significant.

## DISCUSSION

The Muslim Ban Order was a major U.S. policy change impacting the U.S. Refugee Resettlement Program, designed to drastically reduce travel and immigration from Order-targeted nations. After Order issuance, there was an immediate increase in ED visits and ED visits for stress responsive diagnoses among people from Order-targeted nations. Clinic utilization and stress-responsive diagnoses increased before Order issuance, most notably after the 2016 presidential election. The Muslim Ban Order was not a discrete event, as it underwent multiple court challenges and did not go into effect until June 26, 2018. Our findings, even for outcomes which followed similar trends prior to Order issuance, likely reflect the elevated cumulative stress due to multiple restrictive policies and an increasingly hostile climate toward Muslim Americans and Muslim immigrants and refugees in the U.S.^5-7^

Although the estimated differences in utilization and stress-responsive diagnoses are relatively small, they are average differences per person per 30-day period. In a large population, small per-person averages can result in substantial population health changes. In addition to averaging over time, difference estimates may mask heterogeneity in the effect of the Muslim Ban Order on a study population with diverse sub-groups. Our retrospective design and use of EHR data limited our ability to identify different sub-groups, which may respond differently to political stress. Individual-level factors, such as prior trauma, religion, acculturation, and sense of belonging with one’s ethnic group, can influence coping.^22,29^ Factors that may increase susceptibility to stress which are not recorded in EHRs include duration of time in the U.S., whether time spent in a refugee camp, prior interactions with the U.S. immigration and/or refugee administration, employment status, and whether someone is awaiting family reunification.

Estimated effects of the Order may have been attenuated by factors specific to Minneapolis-St. Paul that may not be present in other cities and states. Social capital, ethnic enclaves, and local pro-immigrant policies are important protective factors in Minneapolis-St. Paul that may mitigate the harms from federal policy changes like the Muslim Ban. These local characteristics of Minneapolis-St. Paul have provided social support and promoted civic engagement amongst Somalis living in Minneapolis—paving the way for Minneapolis to elect its first Somali-American City Council member in 2013^30^ and its first Somali-American state legislator Ilhan Omar in 2018.^31^ Social capital, or the ability to secure benefits through social networks and social structures, like community associations or civic organizations, attenuates the negative mental health impacts of self-perceived discrimination^22,25^ through relationships or resources that people mitigate the consequences of prejudice and discrimination.^32^ Ethnic enclaves, or ethnically homogenous social groups, may also protect immigrants from discrimination and related negative health effects. In one study assessing birth weight, residence in a Mexican enclave attenuated risk of low birth weight for Mexican-origin mothers following the 2016 presidential election.^23^ A similar effect was demonstrated when Lauderdale et al’s study on birthweights among Arab Americans after September 11^th^ in California was replicated in Detroit.^8,11^ The initial study demonstrated lower birthweights among Arab Americans following September 11^th^, but this effect was not observed in Detroit, which has a large and strong Arab American community.^11^

Research that aims to understand the impacts of immigration and refugee policy on Muslim immigrants and refugees generally, and Muslim American immigrants and refugees in particular, is limited by the lack of available population-level data. Furthermore, while the increases in healthcare utilization may reflect increased community stress, this study does not directly measure stress nor the relation between healthcare utilization and stress. Further investigations are needed to determine this relation and potential mediators. It is important to consider that healthcare utilization may not be the most sensitive population-level outcome to use as a proxy measure for increased stress in immigrant communities.^15,23^ At baseline, immigrants tend to have lower healthcare utilization compared to people born in the U.S., related to multiple factors including service accessibility, prevalence of chronic illnesses, age, interpreter service availability, cultural health beliefs, and comfort getting care within institutional medical establishments.^23,25^ Furthermore, visiting a medical provider is one of many day-to-day activities, such as school or work attendance, which may be sensitive to acute social stresses and warrant further study.

There are several limitations to this study. First, evaluating the population-level health impacts of the Muslim Ban Order on Muslim Americans is challenging, as most EHR and healthcare data sources do not capture religious affiliation. As such, we used country of origin to estimate Muslim American healthcare utilization.^22,23,29^ Other EHR limitations include data about income and education, which are important factors noted to influence immigrant health care utilization.^33^ Furthermore, when studying a small population, only relatively large effects are easily measured. This may result in an inability to detect small but important effects and precludes nuanced analyses of utilization trends including direct comparison of individuals from Muslim majority nations that were and were not targeted in the Order.

Second, we are limited in our ability to isolate the effects of the Order. Trends for two of our primary outcomes, clinic visits and stress-responsive diagnoses, diverged in the year prior to the Order issuance. Effects which could be estimated through a difference-in-difference analysis may capture the cumulative effect of multiple events taking place around the time of Order issuance and may not reflect its full impact over time. While we are examining changes in utilization around a distinct event, the Order was issued seven days after President Trump’s Inauguration, following a campaign characterized by anti-immigrant and anti-Muslim rhetoric, and it did not go into effect until June 2017. Therefore, we cannot distinguish which specific event caused changes in utilization patterns.

Third, Muslim Americans living in and around Minneapolis-St. Paul are not a homogenous group. Somali Americans, who are racialized as Black, therefore experiencing both racism and anti-Muslim sentiment differently from Arab Muslim Americans racialized as white. This analysis could not differentiate Order impact between these groups, and qualitative methods are better suited to exploring intersectional identities. These and other important differences that are not systematically captured in the EHR could influence the health impacts observed in this study, including: immigration or refugee status, time since immigration, time spent in refugee camps, and acculturation. This study also does not account for second generation immigrants who may experience social and familial stress due to the Order, but be included in the U.S. born comparison group, thereby reducing detected differences between study groups.

Finally, the study group and location are not nationally representative of all groups targeted by the Order, which limits generalizability of our findings. Minneapolis-St. Paul is a large, diverse, urban area with comparatively strong social supports for refugees generally, and Somalis in particular. In other locations with smaller refugee and/or immigration communities and fewer social supports there may be larger negative effects of restrictive immigration and refugee policies.

### Public Health Implications

Increases in primary care utilization prior to issuance of the Order and in ED visits after Order-issuance likely reflect elevated cumulative stress due to multiple factors rather than one particular policy change. Further investigations are needed to evaluate whether increases in utilization are driven by particular subgroups, identify protective factors that convey individual and population-level resilience to political social stressors, and elucidate specific health effects of restrictive immigration policies and cumulative social stress on Muslim Americans.

## Supporting information

STROBE Checklist

## Data Availability

Data available for review upon request.

