## Supplemental Files for "The Impact of the “Muslim Ban” Executive Order on Healthcare Utilization in Minneapolis-St. Paul, Minnesota"

**Table of Contents**

ICD-10 codes used in analysis of stress-responsive diagnoses ………………………………….. Page 2

Analysis of pre-Order trends …………………………………………………….………………. Page 5

Difference in differences estimates for all outcomes ……………………………………………. Page 6

Robustness checks ……………………………………………………………………………….. Page 7

Difference in differences by diagnostic category …………………………………………...…… Page 10

Effects of the Muslim Ban Order on individuals from Muslim-majority

nations not targeted in the Order ……….…………………………………………………………Page 12

**ICD-10 codes used in analysis of stress-responsive diagnoses**

Prior to collecting or observing any data, research team members chose the following ICD-10 codes to include in analysis of stress-responsive diagnoses.

**Table S1: ICD-10 codes included in analysis of stress-responsive clinic diagnoses**

| **Category** | **Diagnoses** | **ICD-10 codes** |
| --- | --- | --- |
| Mental health | Panic disorder  GAD  Anxiety  New SI  Depression  Acute stress disorder  Adjustment disorder | F41.0 Panic disorder [episodic paroxysmal anxiety]  F41.1 Generalized anxiety disorder  F41.3 Other mixed anxiety disorders  F41.8 Other specified anxiety disorders  F41.9 Anxiety disorder, unspecified  F41 Other anxiety disorders (NB)  F51.5 Nightmare disorder  F32 Major depressive disorder, single episode (NB)  F32.0 Major depressive disorder, single episode, mild  F32.1 Major depressive disorder, single episode, moderate  F32.2 Major depressive disorder, single episode, severe without psychotic features  F32.3 Major depressive disorder, single episode, severe with psychotic features  F32.4 Major depressive disorder, single episode, in partial remission  F32.89 Other specified depressive episodes  F32.9 Major depressive disorder, single episode, unspecified  F33 Major depressive disorder, recurrent (NB)  F33.0 Major depressive disorder, recurrent, mild  F33.1 Major depressive disorder, recurrent, moderate  F33.2 Major depressive disorder, recurrent severe without psychotic features  F33.3 Major depressive disorder, recurrent, severe with psychotic symptoms  F33.40 Major depressive disorder, recurrent, in remission, unspecified  F33.41 Major depressive disorder, recurrent, in partial remission  F33.42 Major depressive disorder, recurrent, in full remission  F33.9 Major depressive disorder, recurrent, unspecified  F31.30 Bipolar disorder, current episode depressed, mild or moderate severity, unspecified  F31.9 Bipolar disorder, unspecified  F06.31 Mood disorder due to known physiological condition with depressive features  F43.0 (see below)  F43.1 Post-traumatic stress disorder (PTSD) (NB)  F43.10 Post-traumatic stress disorder, unspecified  F43.11 Post-traumatic stress disorder, acute  F43.12 Post-traumatic stress disorder, chronic  F43.0 Acute stress reaction  F43.8 Other reactions to severe stress  F43.9 Reaction to severe stress, unspecified  R45.7 State of emotional shock and stress, unspecified  R46.6 Undue concern and preoccupation with stressful events  Z63.7 Other stressful life events affecting family and household (NB)  Z63.79 Other stressful life events affecting family and household  Z73.3 Stress, not elsewhere classified  F43.2 Adjustment disorders (NB)  F43.20 Adjustment disorder, unspecified  F43.21 Adjustment disorder with depressed mood  F43.22 Adjustment disorder with anxiety  F43.23 Adjustment disorder with mixed anxiety and depressed mood  F43.24 Adjustment disorder with disturbance of conduct  F43.25 Adjustment disorder with mixed disturbance of emotions and conduct  F43.29 Adjustment disorder with other symptoms |
| Sleep disorders | Insomnia | G47.0 Insomnia (NB)  G47.00 Insomnia, unspecified  G47.09 Other insomnia  F51.0 Insomnia not due to a substance or known physiological condition (NB)  F51.01 Primary insomnia  F51.02 Adjustment insomnia  F51.04 Psychophysiologic insomnia  F51.05 Insomnia due to other mental disorder  F51.09 Other insomnia not due to a substance or known physiological condition  Z72.820 Sleep deprivation  Z72.821 Inadequate sleep hygiene |
| Non-specific | Fatigue  Malaise | R53 Malaise and fatigue (NB)  R53.8 Other malaise and fatigue (NB)  R53.81 Other malaise  R53.83 Other fatigue |
| Gastrointestinal | GERD  PUD (H. pylori)  Abdominal pain (recurrent, NOS, flank pain, epigastric pain)  Irritable bowel | K21 Gastro-esophageal reflux disease  K21.0 Gastro-esophageal reflux disease with esophagitis  K21.9 Gastro-esophageal reflux disease without esophagitis  B96.81 Helicobacter pylori [H. pylori] as the cause of diseases classified elsewhere  K27 Peptic ulcer, site unspecified (NB)  K27.0 Acute peptic ulcer, site unspecified, with hemorrhage  K27.3 Acute peptic ulcer, site unspecified, without hemorrhage or perforation  K25 Gastric ulcer (NB)  K26 Duodenal ulcer (NB)  R10.10 Upper abdominal pain, unspecified  R10.11 Right upper quadrant pain  R10.12 Left upper quadrant pain  R10.13 Epigastric pain  R10.30 Lower abdominal pain, unspecified  R10.31 Right lower quadrant pain  R10.32 Left lower quadrant pain  R10.33 Periumbilical pain  R10.8 Other abdominal pain (NB)  R10.84 Generalized abdominal pain  R10.9 Unspecified abdominal pain  K58 Irritable bowel syndrome (NB)  K58.0 Irritable bowel syndrome with diarrhea  K58.1 Irritable bowel syndrome with constipation  K58.2 Mixed irritable bowel syndrome  K58.8 Other irritable bowel syndrome  K58.9 Irritable bowel syndrome without diarrhea  K59.1 Functional diarrhea |
| Neuro | Headache | G43 Migraine (NB)  G44 Other headache syndromes (NB)  G44.209 Tension Headache  G44.219 Tension Headache, episodic |
| Food-related | Weight loss  Poor appetite | R63.5 Abnormal weight gain  R63.8 Other symptoms and signs concerning food and fluid intake  F50.89 Other specified eating disorder  R63.0 Anorexia  R63.4 Abnormal weight loss  R63.2 Polyphagia  O26.10 Low weight gain in pregnancy |
| Pain Syndromes | Chronic Pain  Back Pain | F45.41 Pain disorder exclusively related to psychological factors  F45.42 Pain disorder with related psychological factors  G89 Pain, not elsewhere classified (NB)  G89.2 Chronic pain, not elsewhere classified (NB)  G89.29 Other chronic pain  G89.4 Chronic pain syndrome  M79.601 Pain in right arm  M79.602 Pain in left arm  M79.603 Pain in arm, unspecified  M79.671 Pain in right foot  M79.672 Pain in left foot  R52 Pain, unspecified  M53.3 Sacrococcygeal disorders, not elsewhere classified M54.2 Cervicalgia M54.41 Lumbago with sciatica, right side M54.42 Lumbago with sciatica, left side M54.5 Low back pain M54.6 Pain in thoracic spine M54.8 Other dorsalgia (NB) M54.89 Other dorsalgia M54.9 Dorsalgia, unspecified |

**Table S2: Emergency Department Acute Stress Diagnoses and Ambulatory Sensitive Conditions**

| **Category** | **Diagnoses** | **ICD-10 codes** |
| --- | --- | --- |
| Acute care stress diagnoses | Acute coronary syndrome  Assault  Suicide attempt  Syncope | I21.0-I21.A; I24.9  Y09., Y08.89x, X99.0xx-X92.9xx; Y01.xxx, Y97.xxx, Y38.3x1- Y38.3x3; T54.1x3-T54.3x3; X98.0xx-X98.9xx; X93.xxx- X95.9xx; Y03.8xx; Y02.0xx, Y00.xxx, Y04.0xx- Y04.2xx, Y04.8xx; Y08.01x- Y08.09x |
| Ambulatory sensitive conditions | Angina  Asthma  Congestive heart failure  COPD  Hypertension  Diabetes complications | I20, I24.0, I24.8–24.9  J45-46  Iii.0, I50, J81  J41-44, J47  I10, I11.9  E10.0–10.8, E11.0–11.8, E12.0–12.8, E13.0–13.8, E14.0–14.8 |

**Analysis of pre-Order trends**

In order to infer effects of the executive order through difference in differences analysis presented in the manuscript, we must assume that in the absence of the executive order, trends in utilization and diagnoses would have been identical across groups. Although we cannot verify this assumption, we can observe whether or not differential trends appear before the executive order is issued. As the visualization in Figure 1 suggests, individuals from Muslim Ban Order targeted nations began to increase clinic utilization, relative to non-Latinx U.S.-born individuals, before the issuance of the executive order. Increases in stress-responsive diagnoses in the clinic are also observed before the executive order is issued. After restricting the comparison group through matching on patient demographics, these pre-Order trend differences are still present.

**Table S3. Analysis of linear time trends in pre-Order periods, Group 1 vs 3 (unweighted)**

|  | Dependent Variable | | | | |
| --- | --- | --- | --- | --- | --- |
|  | Count of Visits or Diagnoses  (Robust standard error) | | | | |
|  | Clinic Visits | Missed Appointments | Clinic Diagnoses | ED Visits | ED Diagnoses |
| Order Targeted | 0.035  (0.005) | 0.017  (0.002) | 0.050  (0.012) | -0.012  (0.002) | -0.065  (0.006) |
| 30 Day Period | -0.0003  (0.00009) | 0.0001  (0.00003) | 0.001  (0.0003) | 0.00003  (0.00005) | 0.002  (0.0002) |
| Order Targeted * 30 Day Period | 0.003  (0.0006) | 0.000006  (0.0002) | 0.005  (0.002) | -0.0002  (0.0002) | -0.002  (0.0008) |
| Constant | 0.158  (0.0007) | 0.029  (0.0003) | 0.420  (0.002) | 0.032  (0.0004) | 0.130  (0.002) |
| Observations (N) | 3016080 | 3016080 | 3016080 | 3016080 | 3016080 |
| R^2^ | 0.00003 | 0.00017 | 0.00001 | 0.00003 | 0.00006 |

**Table S4. Analysis of linear time trends in pre-Order periods, Group 1 vs 3 (after exact matching on demographics)**

|  | Dependent Variable | | | | |
| --- | --- | --- | --- | --- | --- |
|  | Count of Visits or Diagnoses  (Robust standard error) | | | | |
|  | Clinic Visits | Missed Appointments | Clinic Diagnoses | ED Visits | ED Diagnoses |
| Order Targeted | 0.0625  (0.0052) | 0.0042  (0.0023) | 0.1288 (0.0133) | -0.0642  (0.0027) | -0.2404  (0.01) |
| 30 Day Period | -0.0011  (0.0003) | -0.0002  (0.0001) | -0.0023  (0.0008) | -0.0002  (0.0003) | 0.0039  (0.0011) |
| Order Targeted * 30 Day Period | 0.0038 (0.0007) | 0.0003  (0.0003) | 0.0079 (0.0018) | 0.00007 (0.0004) | -0.0039 (0.0013) |
| Constant | 0.1299  (0.0022) | 0.0424  (0.0011) | 0.3395  (0.0059) | 0.0844  (0.0021) | 0.3048  (0.0082) |
| Observations (N) | 1303164 | 1303164 | 1303164 | 1303164 | 1303164 |
| R^2^ | 0.0003 | 0.00001 | 0.00019 | 0.00087 | 0.00078 |

**Difference in differences estimates for all outcomes**

Although we do not ascribe a causal interpretation to all difference in differences estimates, the full set of analyses originally planned are presented in Table S3. This is an expanded version of Table 2 in the main text.

**Table S5. Difference-in-Differences estimates of the effect of the Muslim Ban Order on all outcomes among patients from Order targeted nations (Group 1)**

| **Outcome**  (average per person) | **Difference in Differences Model** | | | | **Matched Difference in Differences Model Estimate**  (SE) | **Generalized Synthetic Control Model Estimate**  (SE) |
| --- | --- | --- | --- | --- | --- | --- |
|  | Means | U.S.-born, non-Latinx | Order-Targeted | **Difference in Differences Estimate**  (SE) |  |  |
| Primary care visits  N = 675,848 | Pre-Order | 0.160 | 0.176 | **0.022**  (0.003) | **0.024**  (0.004) | **0.026**  (0.003) |
|  | Post-Order | 0.164 | 0.202 |  |  |  |
|  | First Difference | **0.004** | **0.026** |  |  |  |
| Missed primary care appointments  N = 152,505 | Pre-Order | 0.029 | 0.046 | **0.002**  (0.001) | **0.003**  (0.002) | **0.002**  (0.001) |
|  | Post-Order | 0.029 | 0.048 |  |  |  |
|  | First Difference | **0.000** | **0.002** |  |  |  |
| Stress-responsive diagnoses in primary care  N = 152,505 | Pre-Order | 0.416 | 0.434 | **0.048**  (0.008) | **0.058**  (0.009) | **0.064**  (0.008) |
|  | Post-Order | 0.432 | 0.498 |  |  |  |
|  | First Difference | **0.016** | **0.064** |  |  |  |
| ED visits per person  N = 112,220 | Pre-Order | 0.032 | 0.021 | **0.004**  (0.001) | **0.005**  (0.002) | **0.004**  (0.003) |
|  | Post-Order | 0.033 | 0.026 |  |  |  |
|  | First Difference | **0.001** | **0.005** |  |  |  |
| ED ambulatory sensitive and acute stress diagnoses  N = 31,861 | Pre-Order | 0.119 | 0.064 | **0.005**  (0.005) | **-0.004**  (0.007) | **0.019**  (0.006) |
|  | Post-Order | 0.133 | 0.083 |  |  |  |
|  | First Difference | **0.014** | **0.019** |  |  |  |

*Note*: As in the main text, effect estimates are additional increases in each outcome (per person per 30-day time period) from the year before to the year after the Order was issued among individuals from Order targeted nations, beyond the increases observed in a reference group. Each outcome is displayed on a separate row. Robust standard errors are included in parentheses for difference in difference estimates with and without demographic matching. Parametric bootstrap standard errors are included in parentheses for generalized synthetical control model estimates.

**Robustness checks**

To assess the robustness of the difference in differences analysis presented in the manuscript, we report two alternative specifications here. First, we report estimates from a model which allows for a linear time trend as an alternative to period fixed effects. Second, we report regression coefficients and sample characteristics for the difference in differences models comparing the individuals from Order targeted nations to U.S.-born non-Latino/a individuals with similar age, sex, race, and insurance to those observed for individuals from Order targeted nations. This reference group was selected using exact matching on all available demographics. Finally, with list the period specific ATT estimates underlying the overall generalized synthetic control ATT estimates. We note, however, that the standard errors on each estimate is imprecisely estimated. Although this analysis attempts to compare individuals from nations targeted by the executive order (Group 1) to U.S.-born non-Latinx individuals with similar demographics and pretreatment trends to those observed in Group 1 (a re-weighted subset of Group 3), it is not entirely successful. Trends continue to differ across these groups after adjustment.

**Table S6. Difference in differences with linear time trend**

|  | Dependent Variable | | | | |
| --- | --- | --- | --- | --- | --- |
|  | Count of Visits or Diagnoses  (Robust standard error) | | | | |
|  | Clinic Visits | Missed Appointments | Clinic Diagnoses | ED Visits | ED Diagnoses |
| Order Targeted | 0.014  (0.002) | 0.017  (0.001) | 0.018  (0.006) | -0.011  (0.0008) | -0.054  (0.003) |
| Order Targeted Post-Order | 0.025  (0.003) | 0.002  (0.001) | 0.048  (0.008) | 0.003  (0.001) | 0.004  (0.005) |
| Period | 0.0001  (0.00003) | 0.00005  (0.00001) | 0.001  (0.00008) | 0.00008  (0.00002) | 0.001  (0.00008) |
| Constant | 0.162  (0.0002) | 0.029  (0.00008) | 0.434  (0.0006) | 0.032  (0.0001) | 0.126  (0.0006) |
| Observations | 6032160 | 6032160 | 6032160 | 6032160 | 6032160 |
| Adjusted R2 | 0.00007 | 0.00018 | 0.00006 | 0.00002 | 0.00007 |

**Table S7. Difference in differences estimates after matching**

|  | Dependent Variable | | | | |
| --- | --- | --- | --- | --- | --- |
|  | Count of Visits or Diagnoses  (Robust standard error) | | | | |
|  | Clinic Visits | Missed Appointments | Clinic Diagnoses | ED Visits | ED Diagnoses |
| Order Targeted | 0.038  (0.002) | 0.002  (0.001) | 0.078  (0.006) | -0.065  (0.001) | -0.215  (0.005) |
| Order Targeted Post-Order | 0.024  (0.004) | 0.003  (0.002) | 0.058  (0.009) | 0.005  (0.002) | -0.004  (0.007) |
| Constant | 0.146  (0.0037) | 0.045  (0.002) | 0.374  (0.010) | 0.084  (0.003) | 0.255  (0.010) |
| Period FE | Yes | Yes | Yes | Yes | Yes |
| Observations | 2606328 | 2606328 | 2606328 | 2606328 | 2606328 |
| Adjusted R2 | 0.00067 | 0.00009 | 0.00056 | 0.0008 | 0.00078 |

**Table S8. Characteristics of HealthPartners patients seeking care in a primary care clinic or emergency department between January 2016 and December 2017, after re-weighting through matching on demographics.**

|  | **Group 1**  **People born in an Order targeted nation**  (n=5,654)  No. (%) | **Re-weighted subset of Group 3**  **U.S.-born, non-Latinx**  (n=102,943)  No. (%) |
| --- | --- | --- |
| **Race** |  |  |
| American Indian/Alaskan Native | 10 (0.2) | 182.1 (0.2) |
| Asian | 44 (0.8) | 801.1 (0.8) |
| Black | 5,229 (92.5) | 95,205 (92.5) |
| Native Hawaiian/Pacific Islander | 5 (0.1) | 91 (0.1) |
| White | 155 (2.7) | 2822.1 (2.7) |
| **Sex** |  |  |
| Female | 3,360 (59.4) | 61,175.9 (59.4) |
| Male | 2,294 (40.6) | 111,786 (40.6) |
| **Age** |  |  |
| 18-24 | 498 (8.8) | 9,227.6 (9) |
| 25-34 | 2,076 (36.7) | 37,448 (20.3) |
| 35-44 | 1,458 (25.8) | 26,587 (25.8) |
| 45-54 | 921 (16.3) | 16,432.5 (16) |
| 55-64 | 518 (9.2) | 9,371.7 (9.1) |
| ≥65 | 403 (7.1) | 7,392.8 (7.2) |
| **Insurance status** |  |  |
| Commercial | 995 (17.6) | 18,116.1 (17.6) |
| Medicare or Medicaid | 4,419 (78.2) | 80,457.2 (78.2) |

*Note:* As in Table 1, missing or unknown data not included in table; sums may not add to 100%

**Table S9. GSC estimates of the effect of the Muslim Ban Order on all outcomes among patients from Order-targeted nations (Group 1) at each time period.**

|  | Dependent Variable | | | | |
| --- | --- | --- | --- | --- | --- |
|  | Count of Visits or Diagnoses  (Parametric bootstrap standard error) | | | | |
| 30-day Period Post EO | Clinic Visits | Missed Appointments | Clinic Diagnoses | ED Visits | ED Diagnoses |
| 0 | 0.034  (0.009) | 0.006  (0.002) | 0.078  (0.016) | -0.002  (0.004) | 0.001  (0.012) |
| 1 | 0.024  (0.006) | 0.0006  (0.003) | 0.058  (0.019) | 0.007  (0.003) | 0.025  (0.025) |
| 2 | 0.036  (0.007) | 0.005  (0.003) | 0.077  (0.022) | 0.007  (0.005) | 0.039  (0.011) |
| 3 | 0.039  (0.008) | 0.009  (0.002) | 0.077  (0.020) | 0.002  (0.007) | 0.018  (0.013) |
| 4 | 0.044  (0.007) | 0.010  (0.003) | 0.103  (0.019) | 0.006  (0.006) | 0.023  (0.014) |
| 5 | -0.006  (0.007) | -0.009  (0.002) | -0.029  (0.021) | 0.004  (0.006) | 0.020  (0.015) |
| 6 | 0.020  (0.006) | 0.0006  (0.002) | 0.047  (0.020) | 0.002  (0.004) | 0.013  (0.008) |
| 7 | 0.038  (0.007) | -0.00006  (0.002) | 0.097  (0.020) | 0.005  (0.004) | 0.020  (0.026) |
| 8 | 0.004  (0.006) | -0.004  (0.003) | 0.024  (0.021) | 0.006  (0.006) | 0.019  (0.025) |
| 9 | 0.054  (0.007) | 0.004  (0.003) | 0.135  (0.022) | -0.004  (0.005) | -0.010  (0.015) |
| 10 | 0.031  (0.006) | -0.0008  (0.003) | 0.081  (0.020) | 0.001  (0.006) | 0.002  (0.019) |
| 11 | 0.026  (0.007) | 0.008  (0.003) | 0.075  (0.020) | 0.009  (0.004) | 0.025  (0.015) |
| 12 | 0.006  (0.007) | 0.005  (0.002) | 0.022  (0.019) | 0.009  (0.008) | 0.039  (0.013) |

*Note:* The ATT effects reported in Table 2 and Table S3 are average effects over 12 30-day periods after the Order was issued. Table entries represent period specific ATT estimates underlying these averages. Parametric bootstrap estimates are based on 1000 replications, but may remain imprecisely estimated.

**Difference in differences by diagnostic category**

The analysis of stress-responsive diagnoses included in the manuscript counts all diagnoses with any of the above codes. Here, we repeat the same analysis separately for diagnoses within each category.

**Table S10. Difference in differences by clinic diagnostic category over 12, 30-day time periods pre- and post-Order**

|  | Dependent Variable | | | | | | |
| --- | --- | --- | --- | --- | --- | --- | --- |
|  | Count of Visits or Diagnoses  (Robust standard error) | | | | | | |
|  | Mental Health | Sleep Disorders | Non-specific | GI | Neuro | Food related | Pain Syndromes |
| Order Targeted | -0.012  (0.0004) | 0.0003  (0.0002) | 0.003  (0.0003) | 0.013  (0.0007) | 0.001  (0.0002) | 0.0008  (0.0002) | 0.012  (0.0009) |
| Order Targeted Post-Order | 0.00008  (0.0006) | 0.0004  (0.0003) | 0.0005  (0.0005) | 0.004  (0.0010) | 0.0006  (0.0003) | 0.0003  (0.0003) | 0.008  (0.001) |
| Constant | 0.018  (0.0003) | 0.003  (0.0001) | 0.003  (0.0001) | 0.010  (0.0002) | 0.002  (0.0001) | 0.001  (0.00007) | 0.017  (0.0004) |
| Period FE | Yes | Yes | Yes | Yes | Yes | Yes | Yes |
| Observations (N) | 6032160 | 6032160 | 6032160 | 6032160 | 6032160 | 6032160 | 6032160 |
| R^2^ | 0.00017 | 0.00002 | 0.0001 | 0.00046 | 0.00004 | 0.00002 | 0.00021 |

**Table S11. Difference in differences by ED diagnostic category over 12, 30-day time periods pre- and post-Order**

|  | Dependent Variable | |
| --- | --- | --- |
|  | Count of Visits or Diagnoses  (Robust standard error) | |
|  | Ambulatory Sensitive | Acute Stress |
| Order Targeted | -0.005  (0.0004) | -0.00041  (0.00011) |
| Order Targeted Post-Order | 0.0007  (0.0007) | 0.00002  (0.0002) |
| Constant | 0.008  (0.0003) | 0.0009  (0.00007) |
| Period FE | Yes | Yes |
| Observations (N) | 6032160 | 6032160 |
| R^2^ | 0.00005 | 0.00001 |

Because some diagnostic trends may be expected to change over different time periods, we additionally include results comparing only the two 30-day periods immediately before and after the Order was issued.

**Table S12. Difference in differences by clinic diagnostic category in the 30 days immediately pre- and post-Order**

|  | Dependent Variable | | | | | | |
| --- | --- | --- | --- | --- | --- | --- | --- |
|  | Count of Visits or Diagnoses  (Robust standard error) | | | | | | |
|  | Mental Health | Sleep Disorders | Non-specific | GI | Neuro | Food related | Pain Syndromes |
| Order Targeted | -0.0146 (0.0011) | 0.00158 (0.0009) | 0.00392 (0.0012) | 0.01922 (0.0026) | 0.00159 (0.0009) | 0.00097 (0.0006) | 0.02019 (0.0034) |
| Post-Order | -0.0018 (0.0005) | -0.00005 (0.0001) | -0.00005 (0.0002) | -0.0004  (0.0003) | -0.00008 (0.0002) | -0.0002 (0.0001) | -0.0011 (0.0005) |
| Order Targeted Post-Order | 0.0021 (0.0018) | -0.0015 (0.0011) | -0.0023 (0.0016) | -0.0014 (0.0036) | -0.0005 (0.0012) | 0.0002 (0.0009) | -0.0015 (0.0048) |
| Constant | 0.0192  (0.0003) | 0.0025  (0.0001) | 0.0035  (0.0001) | 0.0092  (0.0002) | 0.0023  (0.0001) | 0.0013  (0.00008) | 0.0174  (0.0004) |
| Period FE | No | No | No | No | No | No | No |
| Observations (N) | 502680 | 502680 | 502680 | 502680 | 502680 | 502680 | 502680 |
| R^2^ | 0.00018 | 0.00001 | 0.00005 | 0.00065 | 0.00002 | 0.00002 | 0.00028 |

**Table S13. Difference in differences by ED diagnostic category in the 30 days immediately pre- and post-Order**

|  | Dependent Variable | |
| --- | --- | --- |
|  | Count of Visits or Diagnoses  (Robust standard error) | |
|  | Ambulatory Sensitive | Acute Stress |
| Order Targeted | -0.00664  (0.0016) | -0.00074  (0.00019) |
| Post Order | -0.00034  (0.00053) | 0.00011  (0.00011) |
| Order Targeted Post-Order | 0.00687  (0.00329) | 0.00007  (0.00033) |
| Constant | .01052  (0.00039) | 0.00092  (0.00008) |
| Period FE | No | No |
| Observations (N) | 502680 | 502680 |
| R^2^ | 0.00001 | 0.00001 |

**Effects of the Muslim Ban Order on individuals from Muslim-majority nations not targeted in the Order**

Because we observed utilization and diagnosis trends only for 1,254 individuals from other Muslim-majority nations, we are not able to robustly compare the experiences of these individuals from Muslim-majority nations that were and were not named in the executive order. Nonetheless, we note that a difference in differences analysis comparing individuals born in a Muslim majority nation not named in the Order and U.S.-born, non-Latino/a individuals reveals qualitatively similar trends to those observed for individuals from Order-targeted nations.

**Table S14. Difference-in-differences comparing individuals born in a Muslim majority nation not named in the Muslim Ban Order and U.S.-born, non-Latino/a individuals**

|  | Dependent Variable | | | | |
| --- | --- | --- | --- | --- | --- |
|  | Count of Visits or Diagnoses  (Robust standard error) | | | | |
|  | Clinic Visits | Missed Appointments | Clinic Diagnoses | ED Visits | ED Diagnoses |
| Muslim majority | 0.004  (0.005) | 0.004  (0.002) | 0.026  (0.013) | -0.009  (0.002) | -0.035  (0.010) |
| Muslim majority Post-Order | 0.031  (0.007) | 0.007  (0.003) | 0.073  (0.019) | 0.002  (0.003) | -0.002  (0.013) |
| Constant | 0.169  (0.001) | 0.003  (0.0004) | 0.431  (0.003) | 0.003  (0.0006) | 0.108  (0.002) |
| Period FE | Yes | Yes | Yes | Yes | Yes |
| Observations (N) | 5926248 | 5926248 | 5926248 | 5926248 | 5926248 |
| R^2^ | 0.00027 | 0.00007 | 0.00036 | 0.00002 | 0.00006 |

The following nations of origin were included in the set of Muslim-majority nations not named in the Muslim Ban Order (Group 2):

2. Afghanistan
3. Albania
4. Algeria
5. Azerbaijan
6. Bahrain
7. Bangladesh
8. Bosnia-Herzegovina
9. Brunei
10. Burkina Faso
11. Chad
12. Cocos Islands
13. Djibouti
14. Egypt
15. Gambia
16. Guinea
17. Indonesia
18. Jordan
19. Kazakhstan
20. Kosovo
21. Kuwait
22. Kyrgyzstan
23. Lebanon
24. Malaysia
25. Maldives
26. Mali
27. Mauritania
28. Mayotte
29. Morocco
30. Niger
31. Oman
32. Pakistan
33. Palestine
34. Qatar
35. Saudi Arabia
36. Senegal
37. Sierra Leone
38. Tajikistan
39. The Comoros
40. Tunisia
41. Turkey
42. Turkmenistan
43. United Arab Emirates
44. Uzbekistan
45. Western Sahara
